# Interpretable Video-Based Tracking and Quantification of Parkinsonism Clinical Motor States

**DOI:** 10.1101/2023.11.04.23298083

**Authors:** Daniel Deng, Jill L. Ostrem, Vy Nguyen, Daniel D. Cummins, Julia Sun, Anupam Pathak, Simon Little, Reza Abbasi-Asl

**Author notes:** Corresponding authors: Reza Abbasi-Asl, Simon Little.

## Abstract

The ability to quantify motor symptom progression in Parkinson’s disease (PD) patients is crucial for assessing disease progression and for optimizing therapeutic interventions, such as dopaminergic medications and deep brain stimulation. Cumulative and heuristic clinical experience has identified various clinical signs associated with PD severity but these are neither objectively quantifiable or robustly validated. Video-based objective symptom quantification enabled by machine learning (ML) introduces a potential solution. However, video-based diagnostic tools often have implementation challenges due to expensive and inaccessible technology, often requiring multi-camera setups, pristine video collection protocols, or additional sensors that are impractical for conventional use. Additionally, typical “black-box” ML implementations are not tailored to be clinically interpretable, either due to complex and unintuitive algorithms or a lack of analysis on feature stability and optimality. Here, we address these needs by releasing a comprehensive kinematic dataset and developing a novel interpretable video-based framework that accurately predicts high versus low PD motor symptom severity according to MDS-UPDRS Part III metrics. This data driven approach validated and robustly quantified canonical movement features and identified new clinical insights, not previously appreciated as related to clinical severity. Our framework is enabled by retrospective, single-view, seconds-long videos recorded on consumer-grade devices such as smartphones, tablets, and digital cameras, thereby eliminating the requirement for specialized equipment. Following interpretable ML principles, our framework enforces robustness and interpretability by integrating (1) automatic, data-driven kinematic metric evaluation guided by pre-defined digital features of movement, (2) combination of bi-domain (body and hand) kinematic features, and (3) sparsity-inducing and stability-driven ML analysis with simple-to-interpret models. These elements in our design ensure that the proposed framework quantifies clinically meaningful motor features useful for both ML predictions and clinical analysis.

## Introduction

Parkinson’s disease (PD) is a common neurodegenerative disorder characterized by progressive motor symptoms (e.g., bradykinesia, rest tremor, rigidity, postural instability) that can be disabling and significantly impair quality of life^1–3^. The ability to quantify motor symptom progression in PD patients is crucial for assessing and optimizing therapeutic interventions, such as dopaminergic medications and deep brain stimulation (DBS)^4^. Such quantification requires accurate and continual monitoring of motor symptom severity and fluctuations. Currently this objective is only partially satisfied by the *status quo* strategy of intermittent motor assessments assessed at one time point by a single clinician using the Movement Disorder Society-sponsored revision of the Unified Parkinson’s Disease Rating Scale (MDS-UPDRS)^5^. Clinical signs such as finger tapping slowness and decrement (bradykinesia) within the MDS-UPDRS have been discovered by clinical heuristics and codified by expert consensus, precluding well-validated, objective, data-driven and quantifiable assessment of patients. Day-to-day fluctuation of symptoms relies on subjective recall from patients often captured by motor diaries^6^. These approaches are limited by high assessment variance, imperfect recall, and recency bias^7^. To overcome these limitations, the field needs more reliable and objective tracking of PD clinical states.

Technology-based objective symptom quantification, such as those supported by wearable tracking devices and their ability to record kinematic (movement) data, introduces a potential solution^8–10^. Unfortunately, due to device expenses and technical limitations, embodiments of these quantification systems have yet to be regularly adopted into clinical practice for PD assessment^11–13^. More recently, advancements in machine learning (ML) and computer vision have yielded accessible solutions for extracting kinematic information at key anatomical positions from video data without the need for physical marker systems^14–17^. These computer vision solutions have the potential to address the shortcomings of existing methods, significantly enhance diagnostic accuracy, and open new avenues for optimizing personalized medical therapy in PD^18–22^. However, to date, many video-based diagnostic tools have implementation challenges of expensive and inaccessible technology, often requiring multi-camera setups, pristine video collection protocols, or additional sensors that are infeasible for conventional use. Additionally, typical “black-box” ML implementations are not tailored to be clinically interpretable, either due to complex and unintuitive algorithms or a lack of analysis on feature stability and optimality. Therefore, they are generally ineffective in generating novel clinical insights and are challenging to integrate into current clinical care or critically, to develop clinical oversight for. Finally, these tools often focus on prolonged videos from a formal clinical examination or features from a single motor modality, increasing the burden of video acquisition and missing the opportunity to integrate different domains (e.g., body posture, hand movement, facial expression), which has the potential to significantly increase the accuracy and robustness of ML predictions^23–26^. They also typically only predict metrics directly corresponding to a single modality (i.e., predict only MDS-UPDRS finger tapping score) ^22,27^. A truly valuable video-based solution for tracking PD motor symptom progression would need to be affordable, accessible, automated, transparent, and able to obtain rich and clinically relevant metrics for holistic evaluation of PD symptoms^28,29^.

In this study, we address these needs by (1) publicly releasing a comprehensive kinematics dataset from 31 patients all with parkinsonism, and (2) developing a novel video-based framework to automatically predict PD motor symptom high versus low severity according to the MDS-UPDRS Part III metrics (total score). Following interpretable ML principles^30,31^, our primary contribution is to enforce model robustness and interpretability by integrating (1) automatic, data-driven kinematic metric evaluation guided by pre- defined digital features of movement, (2) combination of bi-domain (body and hand) kinematic features, and (3) sparsity-inducing and stability-inducing ML analysis with simple-to-interpret models. We perform a comprehensive kinematic feature stability analysis to identify conserved features across ensemble of models^32–34^ and feature contributions to model outputs via tree SHAP (SHapley Additive exPlanations) analysis^35^. These elements in our design ensure that the model quantifies clinically meaningful motor features providing new clinical insights for quantification of PD severity. Our framework is enabled by retrospective, single-view videos recorded on consumer-grade devices such as smartphones and digital cameras, thereby eliminating the requirement for specialized equipment. In addition, the framework has the advantage of being able to extract rich and meaningful features from just three to seven seconds of video for efficient training and accurate prediction. Validation of features by ML models are enabled by a leave-one-subject-out cross-validation (CV) scheme, an approach mirrored in other PD studies to protect against data leakage and ensure unbiased results^36,37^.

## Results

### Clinical and demographic characteristics

Clinical data were obtained from 31 participants all with parkinsonism and who were evaluated at UCSF as part of a multi-day deep phenotyping cohort study. The protocol included standardized video recordings taken in both “on” and “off” dopaminergic medication states while clinical rating scales were performed. Video kinematic data were retrospectively extracted from the individual subject’s clinical videos. These kinematic data is publicly released as part of this study. The video clips were collected with a single tablet camera in a clinical setting and not initially formalized or collected with optimized settings for a computer vision - ML pipeline. Disease severity was scored using the MDS-UPDRS at the time of assessment. All patients had Parkinsonian symptoms at the time of evaluation. Seven patients were later determined to have Progressive Supranuclear Palsy (PSP) and one patient did not meet clear diagnostic criteria and was classified as PUCS (Parkinsonism of uncertain clinical significance)^38^. Four participants were not taking dopaminergic medication and were only assessed in the “off” medication state.

Patients were dichotomized into two groups associated with low and high Parkinsonian motor symptom severity based on the sample median MDS-UPDRS Part III (motor) score of 32, consistent with literature recommendations^39^. Table 1 summarizes the clinical and demographic characteristics of patients with low (n = 33) and high (n = 25) severity motor symptoms.

**Table 1:** Summary of clinical and demographic characteristics in patients and associated video clips dichotomized by MDS-UPDRS Part III motor score.

|  | Low Motor Impairment (n = 33) | High Motor Impairment (n = 25) | p-value |
| --- | --- | --- | --- |
| Disease Type | PD=33, PSP=0, PUCS=1 | PD=17, PSP=7, PUCS=1 | 0.00<sup>‡</sup> |
| Age (yrs) | 67.1 ± 7.4 | 71.4 ± 6.2 | 0.02* |
| Sex | M=20, F=13 | M=15, F=10 | 0.96<sup>‡</sup> |
| Handedness | R=28, L=3, A=2 | R=21, L=3, A=1 | 0.89<sup>‡</sup> |
| Education (yrs) | 18.3 ± 2.5 | 17.5 ± 2.3 | 0.22* |
| Disease duration (yrs) | 7.4 ± 4.0 | 5.9 ± 3.0 | 0.14* |
| Medication status | On=18, Off=15 | On=9, Off=16 | 0.16<sup>‡</sup> |
| MDS-UPDRS, Part I score | 9.4 ± 5.4 | 10.4 ± 6.3 | 0.55* |
| MDS-UPDRS, Part II score | 8.8 ± 7.5 | 18.4 ± 13.7 | 0.01<sup>†</sup> |
| MDS-UPDRS, Part III score | 18.5 ± 10.3 | 43.4 ± 8.2 | 0.00* |
| Hoehn and Yahr scale | 1.9 ± 0.8 | 3.0 ± 1.1 | 0.00<sup>†</sup> |
| MoCA score | 26.5 ± 3.2 | 23.5 ± 5.0 | 0.01<sup>†</sup> |

PD = Parkinson's disease; PSP = Progressive supranuclear palsy; PUCS=Parkinsonism of uncertain clinical significance; MDS-UPDRS = Movement Disorder Society-Sponsored Revision of the Unified Parkinson's Disease Rating Scale; MDS-UPDRS, Part I = non-motor experiences of daily life; MDS-UPDRS, Part II = motor experiences of daily life; MDS-UPDRS, Part III = motor examination; MoCA = Montreal Cognitive Assessment.
\* Independent 2-sample *t*-test † Mann Whitney *U*-test ‡ Fisher's exact test

The dichotomized groups demonstrated well-balanced characteristics with only age and cognitive profile (MoCA) also showing a difference between the high and the lower severity cohort. The group with high motor symptom severity included patients diagnosed as having PSP, a neurodegenerative disorder with similar Parkinsonian motor symptoms to PD but with more rapid progression^40,41^. Select patients could appear in both high and low severity groups if levodopa medication significantly altered their motor symptom severity to move them from high to low severity. The risks of dependency on hidden covariates and potential data leakage are implicitly addressed during training and validation by ensuring that the data used for validation stemmed from patients unseen in training.

### Automatic extraction of motor features

We designed a computational framework to automatically extract a large array of features representing movement characteristics in raw, unedited video recordings of parkinsonian patients performing motor tasks (Fig. 1a). A small but highly predictive subset of these features was then selected for training and validation of our ML to predict motor symptom severity quantified by MDS-UPDRS Part III metrics (Fig. 1b). Associated with each patient record was a full-body video of walking/gait, a video of the finger-tapping task, or both, collected during the same visit with a standard digital camera. In total, we extracted 40 full-body walking/gait recordings from 25 participants and 48 hand recordings from 27 participants. A comprehensive computer vision pipeline based on deep learning techniques (see **Methods**) was used to extract kinematic time series from each video recording. We extracted kinematic time series from 13 major body landmarks^14^ from each full-body video and kinematic time-series from 8 major hand landmarks from each hand video recording. This was followed by filtering and sub-segmentation of the videos. As a result, all full-length videos were segmented into short, salient, and error-free video segments. From full-body videos, we produced 132 video segments with a mean duration of 5.1 ± 1.1 seconds. From hands videos, we produced 195 video segments with a mean duration of 5.7 ± 1.0 seconds (Supplemental Fig. 2). Removing those with improper or missing associated data entries, we retained 126 full-body video segments and 189 hand video segments. Based on the clinical dichotomization, 65 body video segments and 83 hands video segments were labeled as exhibiting less severe motor symptoms, whereas 61 and 106, respectively, were labeled as exhibiting more severe motor symptoms. The class assignments for the video segments were roughly balanced with marginally higher membership in the “more severe” category.

**Figure 1:**
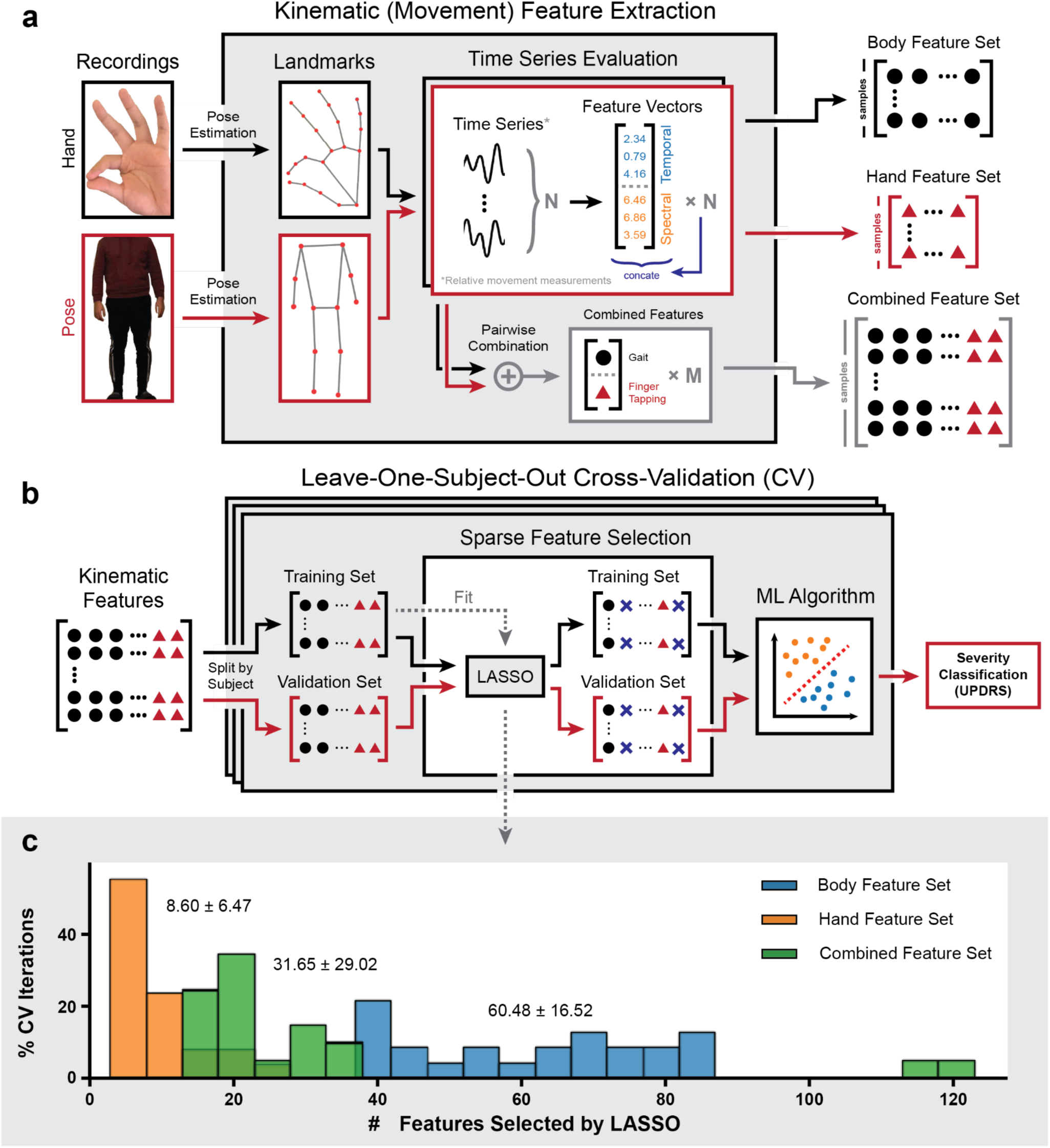
Schematic overview of automatic feature extraction from video recordings for classification of Parkinson’s disease motor symptom severity. **a.** From recordings of participants performing prescribed motor assessment tasks, we extracted movement (kinematic) time series at key landmarks using the pose estimation library MediaPipe. Relative movement measurements were computed based on the extracted signals, from which various temporal and spectral metrics were computed as features. Pairwise combinations of samples from the same patient under the same medication state were performed on the body and hand feature sets to form the bi-modality combination feature set. **b.** To obtain objective measurements of classification performances, we performed a leave-one-subject-out cross-validation (CV), where samples from one patient are held outas the training set for each CV iteration, on each dataset. The CV process is repeated 16 times to account for variabilities in models trained. During CV, a small subset of features with high predictive power with regard to the assigned group labels was selected via least absolute shrinkage and selection operator (LASSO) feature selection. The selected features were used to train the various machine learning (ML) models. **c.** LASSO feature selection consistently identified small sets of salient features during each CV iteration. The average number of features selected at each iteration is reported as mean ± standard deviation.

Relationships between landmarks (*e.g.*, arm-body lateral angle, thumb-index distance) were used to define new relative time series, provided that they were not occluded in the videos or highly similar to another relative time series. The concept behind this design was that some or all of the relative kinematic time series should capture aspects of high-level movement features such as stride patterns or finger- tapping consistency. In total, our framework generated 339 features for each of the 126 video segments from the body and 105 features for each of the 189 video segments from the hands. These features were computed based on the relative time series and their time derivatives with select temporal and spectral kinematic metrics (see **Methods**). Moreover, to unify the motor modalities, we also integrated body and hand feature sets via pairwise concatenation of body and hand feature vectors associated with the same patient with the same medication status. This procedure created 895 combined feature vectors with 496 features. 417 of the combined vectors are labeled as “less severe” and 478 as “more severe”. In all cases, large numbers of salient features were extracted from each video recording without any manual tracking. However, the high-dimensional nature of the feature sets posed a challenge to feature interpretability. To identify a minimal and optimal feature subset for the classification task, we introduced a sparsity-inducing feature selection module based on the least absolute shrinkage and selection operator (LASSO)^42^ technique to the classification framework. This module identified on average 60.48 ± 16.52, 8.60 ± 6.47, and 31.65 ± 29.02 features most important for severity prediction among the body, hand, and combined features, respectively (Fig. 1c). The significantly reduced feature set sizes and enabled further analysis and interpretation of trained models for generating clinically relevant insights.

### Classification of motor symptom severity based on extracted features

To demonstrate that the automatically extracted and sparsified motor features have high predictive power in discriminating between low and high parkinsonian motor symptom severity states, we trained seven different ML models with the generated features (see **Methods**). We then quantified the classification performances using classification accuracy and average area under receiver operating characteristics curve (AUC) scores estimated with inter-patient cross-validation (CV) (Fig. 2; see **Methods**)^43^. In the interest of retaining sufficient sample sizes, we chose to retain data from patients with clinical parkinsonism secondary to PSP and PUCS as well.

**Figure 2:**
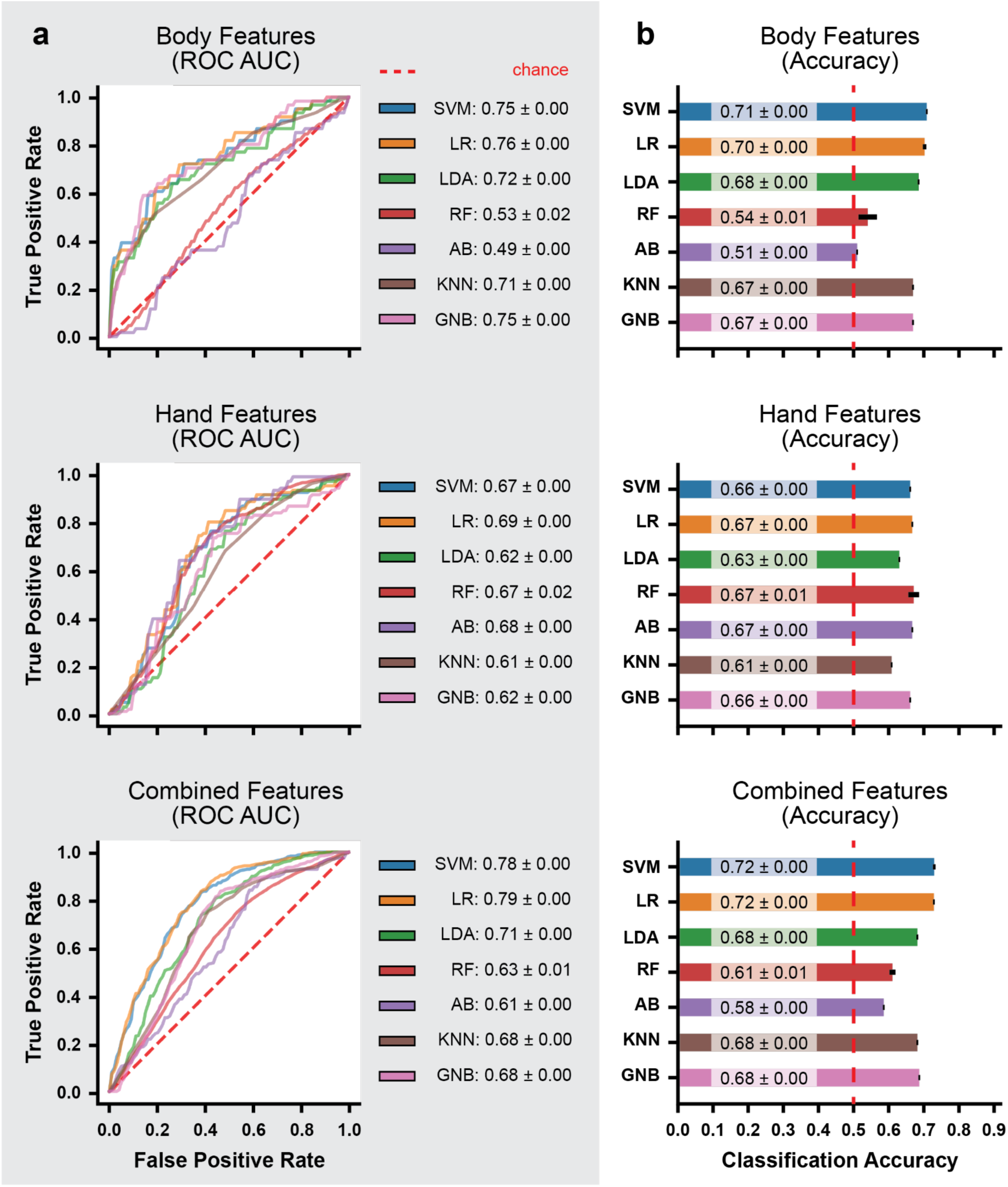
Classification performances of seven selected ML classification models. SVM = support vector machine; LR = logistic regression; LDA = linear discriminant analysis; RF = random forest; AB = adaptive-boosted trees; KNN = K-nearest neighbors; GNB = Gaussian naive Bayes; ROC AUC = area under receiver operating characteristics curve. **a.** Here we show the average ROC curves of the trained ML models during 16-times repeated leave-on-subject-out cross-validation (CV). The corresponding AUC scores are reported in the right margins as mean ± standard deviation. Overall, integrating body and hand features led to improved performances in the most models. **b.** Here we show histograms of the classification accuracies of all models aggregated over all CV iterations. The accuracy scores are reported as mean ± standard deviation at the base of the histogram bars. Similar to the findings when reading the ROC plots, models trained with combined features were more accurate than those trained only on body features, which were more accurate than those trained only on hand features.

When the models were trained and evaluated on the body features, logistic regression (LR), support vector machine (SVM), and Gaussian naive bayes (GNB) classifiers achieved the highest average AUC of 0.76, 0.75, and 0.75, respectively. They were closely followed by linear discriminant analysis (LDA) and k-nearest neighbors (KNN) classifiers, at 0.72 and 0.71 average AUC scores, respectively. The remaining ensemble models, random forest (RF) and adaptive boosted trees (AB), had the chance-level average AUC scores of 0.53 and 0.49. Performance measured by classification accuracy were similar in relative ranking, with SVM and LR achieving the highest average classification accuracy of 71% and 70%, respectively. Lower but comparable classification accuracies of 68%, 67%, and 67% were observed in LDA, KNN, and GBN classifiers, respectively. RF and AB achieved near random accuracies of 54% and 51%. Overall, linear models trained with body features demonstrated decent classification performances. This suggests that the body features, parsed from only a few seconds of walking footage, formed an acceptable representation of the motor characteristics of the corresponding patients.

For the models trained and evaluated using hand features, most classifiers achieved average AUC scores between 0.67 and 0.69, with LR being the most performant at 0.69, closely followed by AB, SVM, and RF at 0.68, 0.67, and 0.67 respectively. LDA and KNN were the least performant at 0.62 and 0.61 AUC. Similarly, most classifiers achieved the average classification accuracy of 66-67%, with the exception of LDA and KNN at 63% and 61%, respectively. Similarly, overall, the classification performances of most models trained with hand features were slightly poorer compared to models trained with body features. This may be explained by the fact that movement characteristics measurable by the finger-tapping task are specific and limited, whereas whole-body motor assessments, such as the walking task used in this investigation, may contain richer information for diagnosing the overall level of motor impairment.

Integrating both body and hand features into a single model sustained or improved the classification performances. SVM and LR achieved the highest average AUC scores at 0.78 and 0.79, outperforming the single modality models. LDA, KNN, and GNB achieved lower AUC scores of 0.71, 0.68, and 0.68, consistent with or slightly lower than their counterparts trained with only body or hand features. RF and AB achieved the lowest scores of 0.63 and 0.61. In terms of classification accuracy, SVM and LR achieved an average accuracy of 72%, with LDA, KNN, and GNB following closely at 68%. The least performant RF and AB classifiers achieved average accuracies of 61% and 58%, which still offered an improved lower bound on the accuracies compared to training only with body features. Overall, integrating body and hand features led to improved performances in the most accurate models and similar performances in the remaining models.

### Feature stability analysis and clinical insights

The classification performances demonstrated that our framework is capable of extracting optimal features for discriminating low and high motor symptom severity. However, direct interpretation of trained ML models is usually challenging due to variance in LASSO feature selection. The variance was a consequence of distinct data partitioning during CV. To allow insight into the most important features and their contributions to model outputs, we performed ensemble feature stability analysis, aggregating selection counts of features over all leave-one-subject-out CV iterations for each type of classifier model. Specifically, we considered features selected in at least 50% of all iterations as stable based on the ensemble paradigm^32^, which is a trade-off between retaining all potentially relevant features and retaining only stable features to facilitate interpretation

Performing the analysis on the most performant combined feature set, we identified 9 body and 5 hand features with high stability for interpretation. For all identified features, there exist statistically significant differences (p ≪ 0.05; independent 2-sample *t*-test) in group means between the low and high motor symptom severity groups (Fig. 3a). Visualizations of the landmark relationships from which metrics were derived can be seen in Fig. 3c.

**Figure 3:**
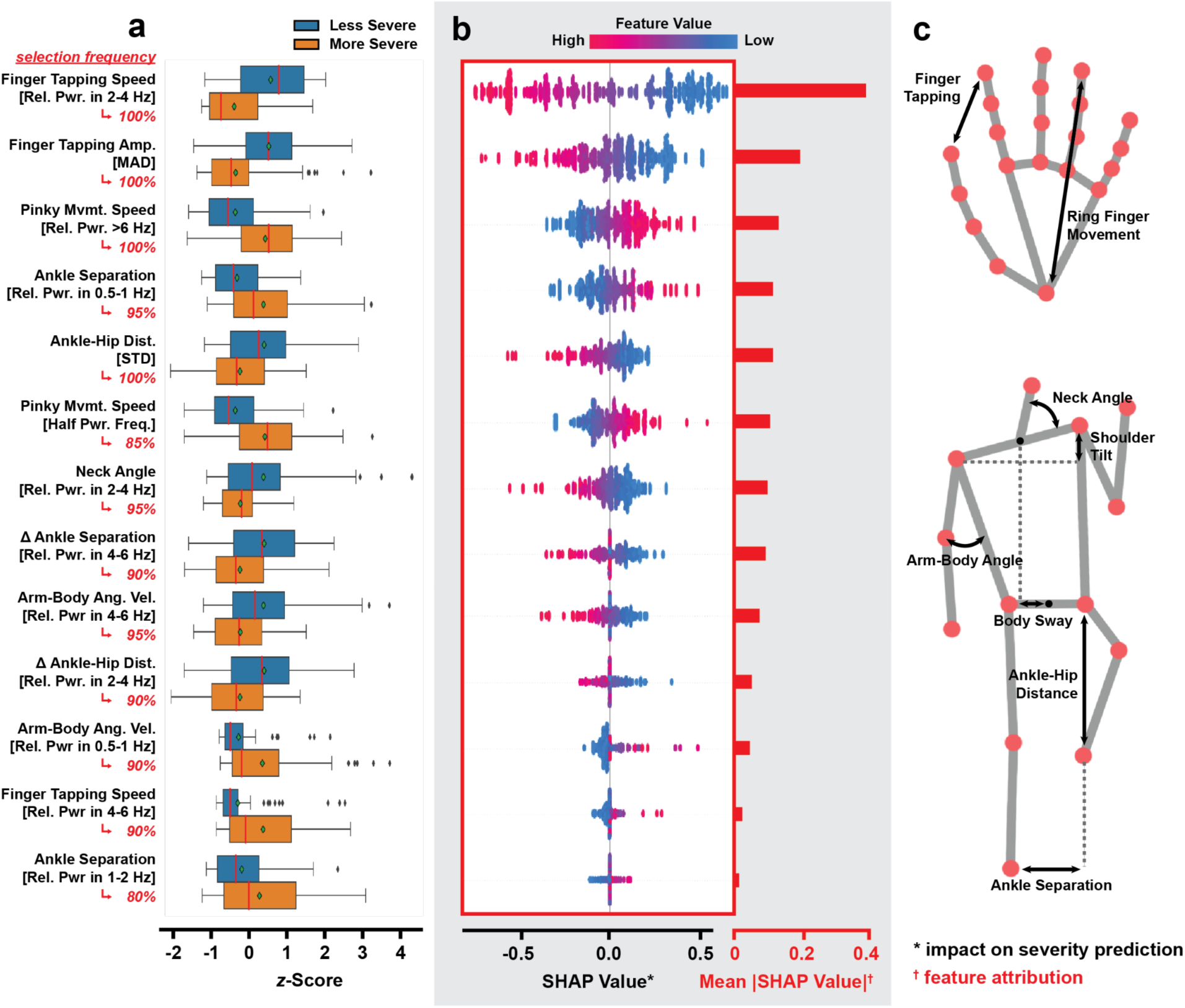
Group differences in stable features and their contribution to ML model predictions. STD = standard deviation. MAD = median absolute deviation. The stable features were chosen according to the criteria that they must be selected in at least 50% of all models trained during cross-validation (CV). **a.** Point plot showing the statistically significant differences (p ≪ 0.01) in group means for the selected features between groups with low and high levels of motor impairment. The feature values are *z*-score normalized. **b.** The swarm plots on the left show individual SHAP values of feature values encountered during CV training of a logistic regression (LR) classifier. The histograms on the right show the mean absolute SHAP values of each feature. A higher SHAP value indicates a stronger bias toward predicting the “less severe” class label. A higher mean absolute SHAP value for each feature corresponds to its level of feature attribution in relation to the predicted label. **c.** Visualizations of landmark relationships from which kinematic metrics were derived. The body and hand skeletons also show important landmarks and other landmark relationships were based on in our extraction pipeline.

The disparities between feature values in the dichotomized groups and their impact on deciding the severity state of patients are compatible with the current clinical understanding of PD and parkinsonism symptoms. Specifically, in patients with higher symptom severity, the model pulled out higher variability in the inter-ankle distance during walking, suggestive of unsteadiness. Additionally, higher movement of the pinkie finger was seen during the finger tapping task which is strictly meant to be restricted to index finger movements only. This may reflect an increase in rigidity or loss of movement discriminability between fingers in more severe clinical Parkinsonian cases. More severe cases also had lower hip-ankle variability during gait, reflective of hip flexion reductions and gait “shuffling”. Increased axial rigidity was supported by reduced movement in the neck (relative power in the 2-4 Hz of neck angle) during gait as well as reductions in finger tapping as measured by the finger tapping speed and amplitude.

We also found that the power spectral density for arm-body lateral angular velocity has a distribution shift with the severity of the motor symptoms. Specifically, the relative power in 4-6 Hz of arm-body lateral angular velocity is lower for patients with more severe motor symptoms, while the relative power in 0.5-1 Hz is lower for patients with less severe symptoms (see also Supplemental Fig. 4). This suggests that with more severe PD motor symptoms, patients have increased lower-frequency changes and reduced higher-frequency changes in arm body lateral angle, suggestive of axial rigidity.

Similarly, there is a distribution shift for the power spectral density of finger tapping speed. Patients with more severe symptoms have higher relative power in 4-6 Hz of their finger tapping speed suggesting jerky / sudden interruptions to regular tapping as this frequency range is higher than the actual tapping frequency. The relative power in 2-4 Hz of the finger tapping speed decreases with severity consistent with incomplete finger tapping for patients with more severe motor symptoms.

The directions of the group differences were consistent with the SHAP values, which measure the impact feature values have on model outputs, estimated with a separate LR classifier using all 14 features (Fig. 3b). In this case, a higher SHAP value biases the model towards assigning a sample the “less severe” label. Features whose values have a positive correlation with the estimated SHAP values were consistent with features with higher group means for participants with less severe motor symptoms. Moreover, based on the mean absolute SHAP value of each feature, which reflects overall feature attribution, body features had the most predictive power, higher than hand features associated with finger tapping.

To further validate the optimality of the selected stable features, we compared the 2D projection results of each feature set (body, hand, combined) with and without stable feature selection (Fig 4). Here, the projection algorithm used was MDS (multidimensional scaling), which tends to preserve the global structures present in the data. When all features generated by our framework were retained prior to projection, the final class clusters were poorly separated in all three cases (Fig. 4a). In contrast, the class clusters saw improved linear separability when only stable features were retained prior to projection.

**Figure 4:**
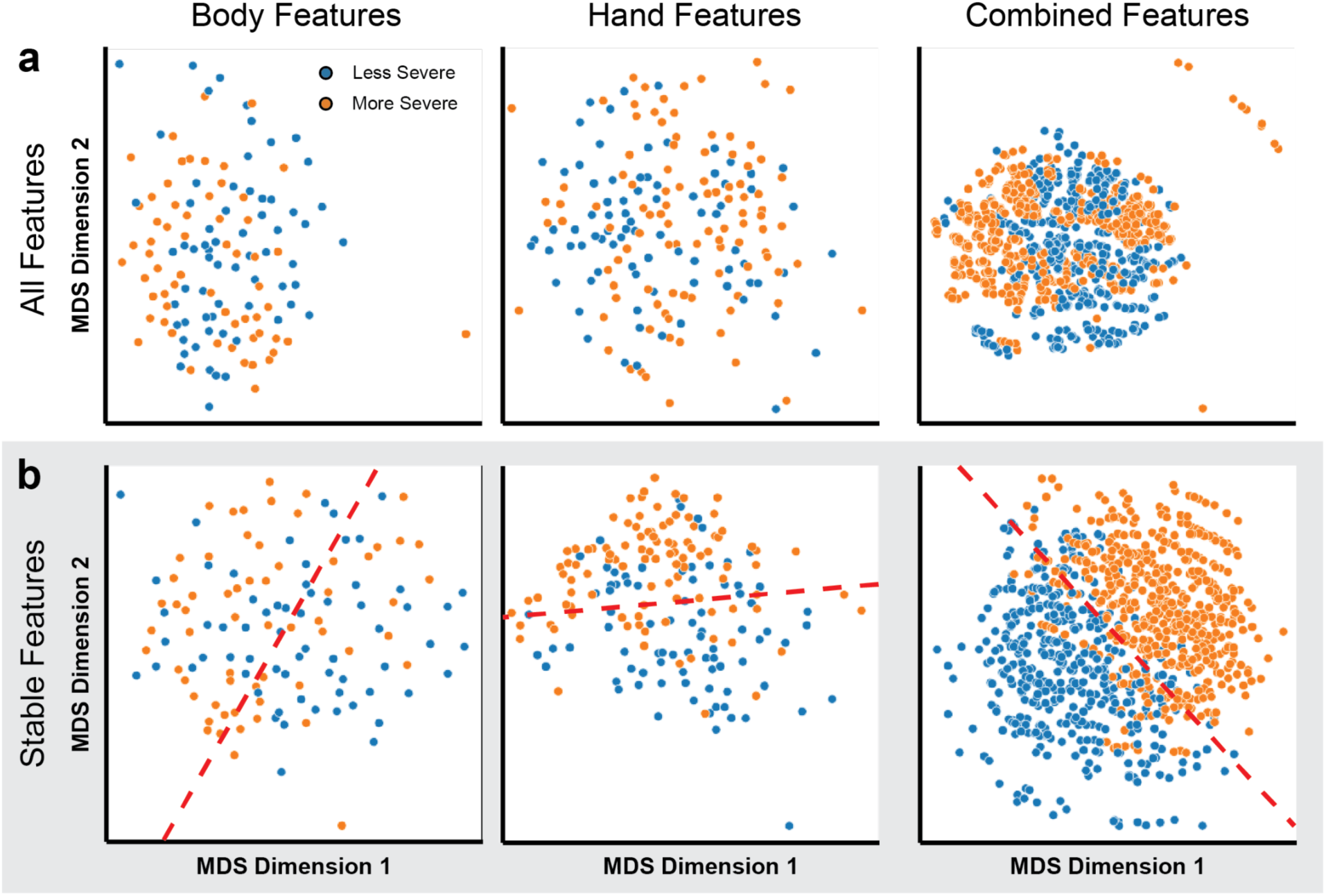
Scatter plots comparing 2D projections of datasets with and without stable feature selection. The projections were computed using MDS (multidimensional scaling). **a.** Poor separability of class clusters in 2D projection space when all features in the datasets were used. **b.** Improved separation (especially when both body and hand features were included) of class clusters when only stable features were used.

Samples containing body or combined features experienced the most significant increase in separability, consistent with the high classification performance in models trained with them. (Fig. 4b). These results suggest that the stable features identified through ensemble analysis well-characterized the low and high motor impairment groups and are optimal for the classification task at hand. In conclusion, this form of ensemble analysis not only unified the CV iterations and increased model interpretability but also helped to identify and preserve the most important features.

## Discussion

Our study demonstrates that an interpretable computer vision - ML pipeline is able to accurately classify parkinsonian motor severity (high versus low) using very brief video recordings of gait and finger tapping. The classification accuracy was significantly improved using a combined pipeline that incorporated both walking and finger movements. The combined model classification was comparable to previous studies that have relied upon complex and extensive hardware solutions including wearable sensors and prolonged, formalized clinical videos. Notably, our study was performed using only very short video clips from a retrospective clinical library; video recordings were not performed in a laboratory under standardized conditions nor optimized for computer vision or machine learning. This supports translation and scalability to real-world applications both in-clinic and potentially extended to outside the clinic.

Clinical movement disorder diagnosis and tracking has shown limited fundamental conceptual advancement over the course of the last 200 years, relying mainly on subjective (expert) recognition and classification of symptoms from visual inspection. This is unsurprisingly significantly limited by inter-rater variability and lack of reproducibility. However, in addition to providing an accurate, objective, evaluation of Parkinsonian motor severity, unlike many ML schemes, our algorithm has the potential to provide, rather than obscure, clinical insight, through bespoke feature stability analysis. Here the algorithm identified a number of classic parkinsonian features including finger tapping speed and arm swing during gait which serve as useful validation. Moreover, our pipeline identified other features including pinky finger stability during the finger tapping task and neck angle features that appear to drive classification and might serve as useful features for clinical classification, that may to date have been underappreciated in medical training. A bidirectional relationship between clinical expertise and ML-led feature identification has the potential to improve classification accuracies and clinical knowledge.

Here our algorithm was used to classify high versus low parkinsonian severity instead of predicting MDS-UPDRS scores directly with regressive models due to limited data availability. In the future, with larger datasets this could be further trained to provide objective, quantitative, diagnostic evaluations that could support classical clinical diagnostic pathways. In addition to single time point evaluations, this approach could in principle be extended to at-home implementation. This has the potential to support chronic symptom tracking in order to monitor naturalistic motor fluctuations over time toward personalized optimization of therapy. Online pose estimation (with immediate deletion of raw video footage) plus automated person recognition have the potential to address potential privacy and security concerns.

The accuracies reported in our study are not directly comparable to existing studies which reported relatively higher classification accuracy differentiating PD from healthy control using movement data (average accuracy of 89.1 ± 8.3%)^44^ for three primary reasons: First, our study classifies severity in patients diagnosed with Parkinsonian symptoms (rather than against healthy controls) and attempts to predict high versus low total MDS-UPDRS Part III score. This is a more difficult task compared to the classification of finger tapping or gait scores, which is primarily used in past studies. Second, our approach is designed to use short (∼5) second-long video segments captured by consumer-grade devices such as smartphones, tablets, simple digital cameras. Previous studies, on the other hand, benefit from the movement data collected using a variety of camera-based, sensor-based, or other miscellaneous recording devices captured over multi-minute long recordings. Therefore, they lacked the technical simplicity offered by our framework^45^. Third, we prioritize model interpretability over performance to determine highly stable and predictive features of hand and body, providing reliable clinical insights.

There are a number of limitations to the present study. First, the relatively small cohort size might limit or bias the model performances reported. A larger cohort of patients is required to fully validate the framework’s reported performance and objectivity, particularly with respect to important features identified. Ideally, the cohort should contain more diverse subjects, with patients at all stages of disease progression and with healthy controls. Having a larger cohort could also enable fine-grain analysis such as regression on UPDRS scores to validate the framework’s utility in obtaining detailed clinical diagnoses. Second, the performances reported may be affected by the quality of the video data, which was not collected in a manner specifically designed and optimized for a computer vision task. This has resulted in some suboptimal data unsuitable for ML analysis. Standardization of video acquisition and analysis protocols will be beneficial for the framework’s objective assessment of PD symptoms, as well as adaptability and scalability. However, this also demonstrates the robustness of our technique to real-world clinical application. Additional pre-processing should also be explored to increase the pose position estimate validity, including evaluation of a wider suite of available pose estimation computer vision software packages. However, this dataset represents a floor to classification accuracy. Finally, we chose to perform stability analysis based on selection frequency by LASSO during training to promote feature interpretability. However, the method might be overly conservative and ignorant of some important features, especially those with collinearity. Future developments could consider methods such as creating “proto-features” from clusters of correlated features^46,47^, which might better preserve important features.

In conclusion, our framework effectively expands upon previous research in PD quantification and addresses many of the shortcomings for a simple yet comprehensive video-based solution. Analyzing a cohort of parkinsonism patients with the proposed framework, we showed that our approach extracted and identified salient kinematic features that could be used to train accurate ML models for predicting low- and high-severity states for motor impairment with high accuracy. Follow-up studies should focus on further refining the framework, increasing the degree of automation, and validating it in larger, representative cohorts. The framework should also be extended to incorporate additional motor modalities, such as facial expressions and speech, as well as non-motor modalities such as neurological data. Future directions should also include exploring the framework’s utility in predicting other clinically relevant outcomes in PD and in application to other neurological movement disorders such as dystonia and essential tremor.

## Acknowledgements

The authors would like to thank Chirag Sharma, Russell Ro, Ian Bledsoe, Ethan Brown, Howie Rosen, Carlie Tanner, Michael Geschwind, Kevin Lieu, and Bruce Miller for their comments and contributions to the recruitment and evaluation of the patients. RA, SL, and JLO would like to acknowledge support from the UCSF Innovation Ventures.

## Data availability

We have publicly released the kinematic data from 31 participants with Parkinsonism. The data is freely available at https://github.com/abbasilab/Video-Tracking-PD.

## Code availability

The software package is freely available at https://github.com/abbasilab/Video-Tracking-PD.

## Competing Interests

The authors declare no competing interests.

## Methods

### Participants and assessment of motor symptoms

A cohort of patients presenting with parkinsonism symptoms, including idiopathic PD and PSP, was recruited at the UCSF Movement Disorder and Neuromodulation Center. Qualitative and Quantitative assessments of motor and non-motor symptoms of participating patients were conducted by a movement disorders neurologist based on MDS-UPDRS, the H&Y scale, and the MoCA scale. The main assessment metric used in this study for measuring motor symptom severity was the MDS-UPDRS Part III score ^22,48^. The MDD-UPDRS uses an ordinal scale ranging from 0 to 132, where higher values indicate greater motor impairment. Motor assessments using MDS-UPDRS Part III were conducted for the “off” and “on” dopaminergic medication states. For the “off” state, participants were withdrawn from their medications for a minimum of twelve hours prior to assessment. For the “on” state, participants were given a standard morning dose of Levodopa medication and evaluated one hour later. Not all patients were prescribed dopaminergic medication, and thus some were only assessed in the “off” state. Concurrently with the clinical assessments, video recordings of participants were captured with a consumer-grade iPad tablet recording device at full HD (1920x1080) resolution and 30 frames per second, mounted on a tripod in the clinical care area. During recording, participants were instructed to perform finger-tapping and gait tasks as described in MDS-UPDRS sections 3.4 and 3.10. For the finger-tapping task, participants were instructed to tap their index finger on their thumb a minimum of ten times with maximally achievable speed and amplitude. For the gait task, participants were instructed to walk toward and away from the camera for a minimum of 10 meters (30 feet) each way. The resultant videos underwent no additional editing. All participants provided written consent forms for the use of personal health information in research and release. Privacy and confidentiality protection have been explicitly addressed with IRB approval. The raw videos were not authorized for release.

### Pose estimation and signal processing

For each video, kinematic (movement) time series at select body and hand landmarks (Fig. 5) were extracted using the Pose and Hands tracking solutions from MediaPipe^14^. Mediapipe is an open-sourced framework for building multimodal machine learning pipelines. It is cross-platform (server, iOS, Android) and uses a graph-based pipeline to perform processing and inference functions on multimodal input streams, such as vision and audio. Through this framework, we employ pre-trained hand and pose models, which were trained on ∼30k and 85K (25k of which performing fitness exercises), respectively. All of these images were annotated by humans^49^. The hand model infers 21 3D landmarks of a hand from a single frame, while the pose model predicts the location of 33 pose landmarks. The two-stage pipeline used by MediaPipe for pose and hand tracking consists of (1) an autoencoder detector, similar to feature pyramid networks, for finding bounding box for pose or hand ^50^, and (2) an convolutional encoder tracker for landmark localization informed by the bounding box. While current state-of-the-art approaches rely primarily on powerful desktop environments for inference, Mediapipe pose and hand models achieve real-time performance on mobile phones. MediaPipe, as well as alternatives such as DeepLabCut and OpenPose, have been adopted for accurate extraction of kinematic data from video recordings for clinical analysis of PD^24,51–53^.

**Figure 5:**
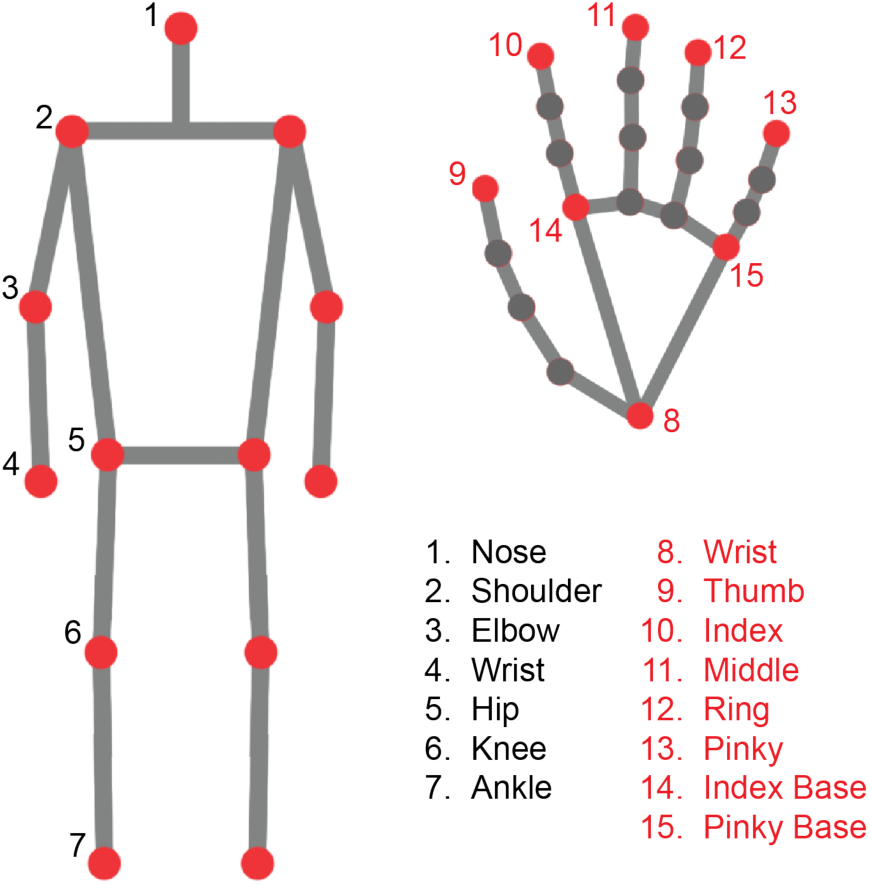
Body and hand skeletons with key landmarks used in analysis labeled. For simplicity, we only labeled relevant landmarks on one side of the body and one of the hands. Some labels were renamed and differed from the official nomenclatures provided by MediaPipe.

Moderate Gaussian smoothing was applied to the extracted signals to reduce random fluctuations due to tracking inaccuracies, implemented as a weighted sum over a 5-point rolling Gaussian window (σ = 0.5) with the python data manipulation library pandas (version 1.4.0)^54^. Time points at which major tracking errors in any time series, *e.g.* missing data, invalid numerics, severe flickering, occurred were identified and marked for removal. Specifically, severe flickering was detected by checking for rapid zero-crossing in distance between shoulders. Time points at which specific posing requirements were not met were also marked for removal. For body recordings, the requirements were that the subject being filmed must be fully standing and roughly facing forward or backward relative to the camera. We determine if a subject is standing by verifying that the apparent lower leg length (ankle-knee distance) is at most twice the apparent upper leg length (hip-knee distance). We approximately determine if a subject is facing forward or backward by verifying that the horizontal distances of left-wrist-to-left-hip and right-wrist-to-right-hip are of opposite signs, *i.e.* the hands are on opposite sides of the body. For hand recordings, the requirement was that one and only one hand must be raised (and assumed to be engaged in active finger-tapping). We determine this by verifying that the vertical distances of left and right index-finger-to-wrist are of opposite signs, *i.e.* one hand is pointed up and the other down. For each invalid time point identified, a time window within 0.333 seconds (10 time points) from the invalid point was removed from all extracted time series.

As a byproduct, the filtering operation effectively segmented the movement data, resulting in sets of short time series at key landmarks grouped by video segments. To equalize the video segments with respect to duration (and by extension the amount of contained information), segments with a duration less than 3 seconds were discarded and segments with a duration greater than 8 seconds were further sub-segmented. The minimum and maximum duration values were empirically chosen such that each segment contained sufficient kinematic information, *i.e.* multiple stride or finger-tapping cycles, for non-trivial extraction of motor features while maximizing the number of samples available for subsequent model training and classification.

### Computing relative movement measurements

Similar to strategies seen in existing literature^53^, we generated relational time series from the filtered and segmented kinematic time series based on predefined interactions between two or more landmarks.

Unlike previous studies where the relationships are manually selected, we included most relationships between landmarks as long as they are not occluded or invalidated by the retrospective view of the videos or have significant overlap with another relationship. These new time series collectively captured aspects of healthy and pathological movement features without explicit computation of the movement features. The list of time series computed is provided here:

1. Neck angle (angle between nose, mid-shoulder, and left shoulder)
2. Arm-body lateral angle (angle between elbow, shoulder, and hip, lateral to body)
3. Left/right wrist-shoulder distance
4. Left/right ankle-hip distance
5. Ankle separation (horizontal displacement between ankles)
6. Knee separation (horizontal displacement between knees)
7. Body sway (horizontal displacement between mid-shoulder and mid-hip)
8. Leg raise (vertical displacement between ankles)
9. Shoulder tilt (vertical displacement between shoulders)
10. Hip tilt (vertical displacement between hips)
11. Finger tapping (distance between thumb and index finger of active hand)
12. Middle finger movement (distance between middle finger and wrist of active hand)
13. Ring finger movement (distance between ring finger and wrist of active hand)
14. Pinky finger movement (distance between pinky finger and wrist of active hand)

Note that the filtering step ensured that only one hand will be actively performing finger-tapping for any given time segment; therefore, distinguishing between left and right-sided hand movements was unnecessary as the resting hand will necessarily have little to no movement and could have its time series discarded.

### Time series assessment with kinematic metrics

Once we computed the relational time series, as the apparent body and hand sizes change depending on the distance between the subject and the camera, we normalized the new time series at each time point to equalize the scale and allow direct comparison. For body signals, normalization was achieved by dividing the time series by the body length (distance between mid-shoulder and mid-hip) and centered mid-hip. For hand signals, normalization was achieved by dividing the time series by the palm length (distance between wrist and midpoint between bases of index and pinky fingers) and centering at the wrist. The normalization was followed by kinematic metric evaluation of each time series, during which various temporal (distribution-based) and spectral (spectral density-based) metrics were used to provide a statistical summary of the body and hand movement characteristics (Table 2). This process was repeated for the derivatives of the relational time series, calculated as the rate of change between adjacent time points.

**Table 2:**
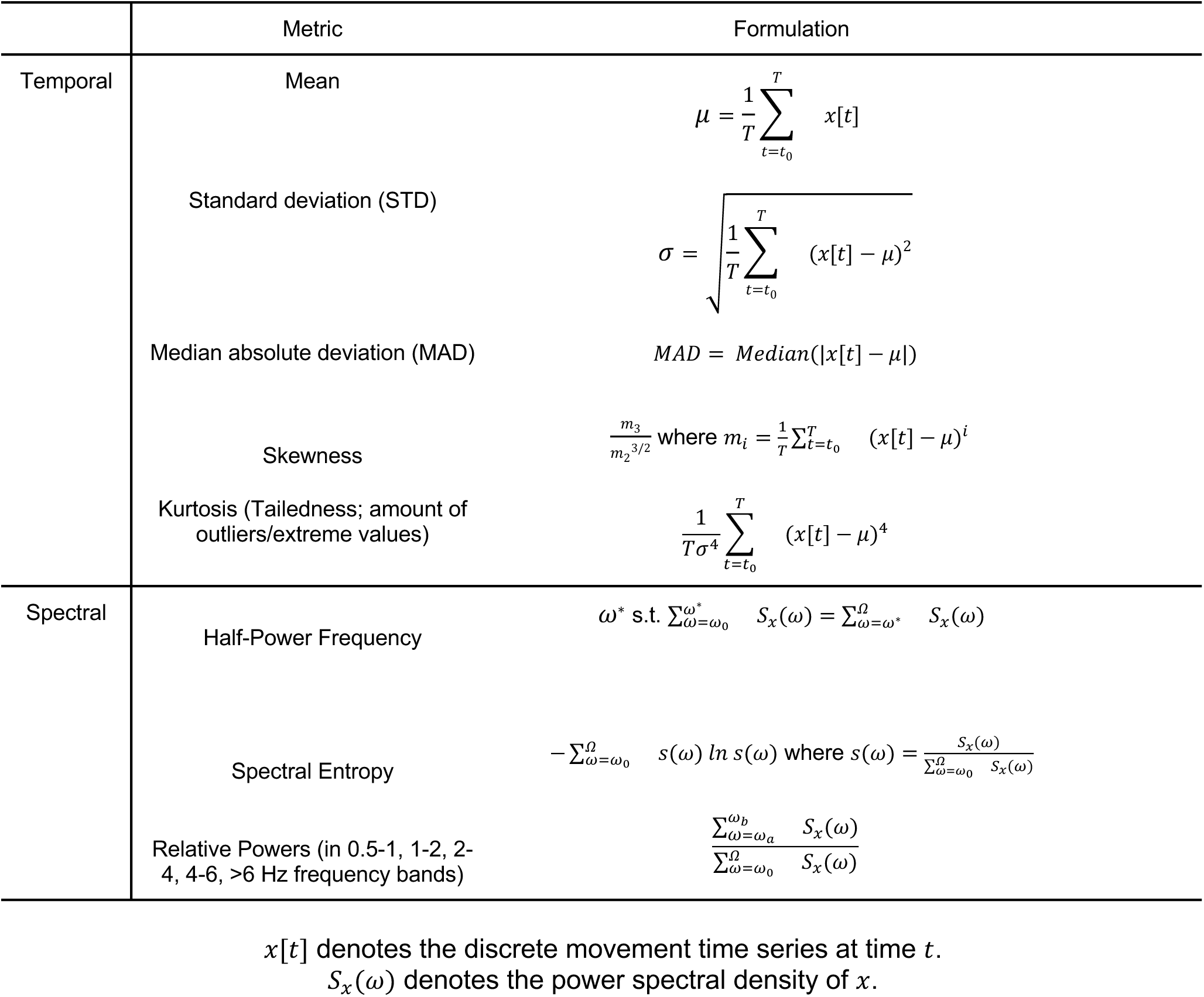
Temporal and spectral metrics used to characterize movement measurements extracted from video recordings.

The computed metric values formed the feature vectors associated with each time segment and were used in subsequent ML training and classification. Since only one hand is performing active finger-tapping at any time in the videos, features from the non-active hand are discarded. Considering the common unilateral development of PD symptoms, we needed to retain both left and right-sided body features. This posed a challenge to compare features between subjects whose manifestation of unilateral features might be on opposite sides of the body. To address this, we recategorized left and right-sided features of the same type as “minimum” and “maximum” features by ranking numerically, thereby eliminating the sidedness of features. In addition, to unify the body and hand kinematic features and increase classification performance, we generated combined feature vectors from all valid combinations of body and hand feature vectors, so long as they were associated with the same patient record, *i.e.* same patient under the same medication status.

### Feature selection and classification

The scikit-learn package (version 1.1.2) in Python was used for feature selection and classification on low and high motor symptom severity for all three feature sets generated by our framework. To generate training and validation data in a robust manner, we conducted the aforementioned procedure with a custom leave-one-subject-out CV, repeated 16 times to account for model variability due to hyperparameter tuning. The leave-one-subject-out protocol selects a different patient at each CV iteration and isolates all samples related to that patient as the testing set. This form of CV is necessary to prevent dependency on hidden confounding variables and thus data leakage. Once partitioned, the training and validation sets were centered and normalized feature-wise based on means and standard deviations from the training set. Then, to reduce feature dimensionality, address multicollinearity, and improve interpretability, we applied to all feature sets as a pre-training step LASSO feature selection, which is an L1-regularized, sparsity-inducing algorithm. Finally, classification performances were evaluated using seven different ML algorithms: (1) linear discriminant analysis (LDA)^55^; (2) logistic regression (LR)^56^; (3) support vector machine (SVM)^57^; (4) random forest (RF)^58^; (5) adaptive-boosted trees (AdaBoost; AB)^59^; (6) K-nearest neighbors (KNN)^60^; and, (7) Gaussian naive Bayes (GNB)^61^. The regularization parameter for LASSO feature sparsity, as well as relevant hyperparameters for each classification model, were chosen via hyperparameter optimization with nested leave-one-subject-out CV during each CV iteration. Classification accuracies and AUC scores were recorded for each classifier, where accuracy measures the model’s ability to classify on the defined class labels and AUC measures the model’s sensitivity and generalizability by looking at all probabilities for assigning labels. Accuracy and AUC scores during CV were aggregated and reported as mean ± standard deviation.

### Feature stability analysis

To address the challenge of interpreting membership variance in salient feature subsets selected via LASSO during CV, we performed feature stability analysis based on frequencies of selection. Here, we defined a feature as being stable if it was selected by LASSO in at least 50% of all CV iterations. Once the stable features have been identified, their contributions to model predictions were evaluated with SHAP analysis on the most performant model using the python package SHAP (version 0.40.0).

Additional validation was performed by comparing the 2D projections of the datasets with and without stable feature selection. The algorithm used for projection was multidimensional scaling (MDS), an unsupervised dimensionality reduction algorithm that preserves global structures in the data.

### Statistical Analysis

Continuous variables were presented as mean ± standard deviation and compared between low and high motor scoring groups with independent 2-sample *t*-test if normally distributed or with the Mann-Whitney *U*-test if otherwise. Categorical variables were presented as counts and compared between scoring groups with Fisher’s exact test, which is analogous to the Chi-squared test but suitable for small-sized samples. A *p*-value less than 0.05 was considered statistically significant.

## Supplemental Materials

**Supplemental Figure 1:**
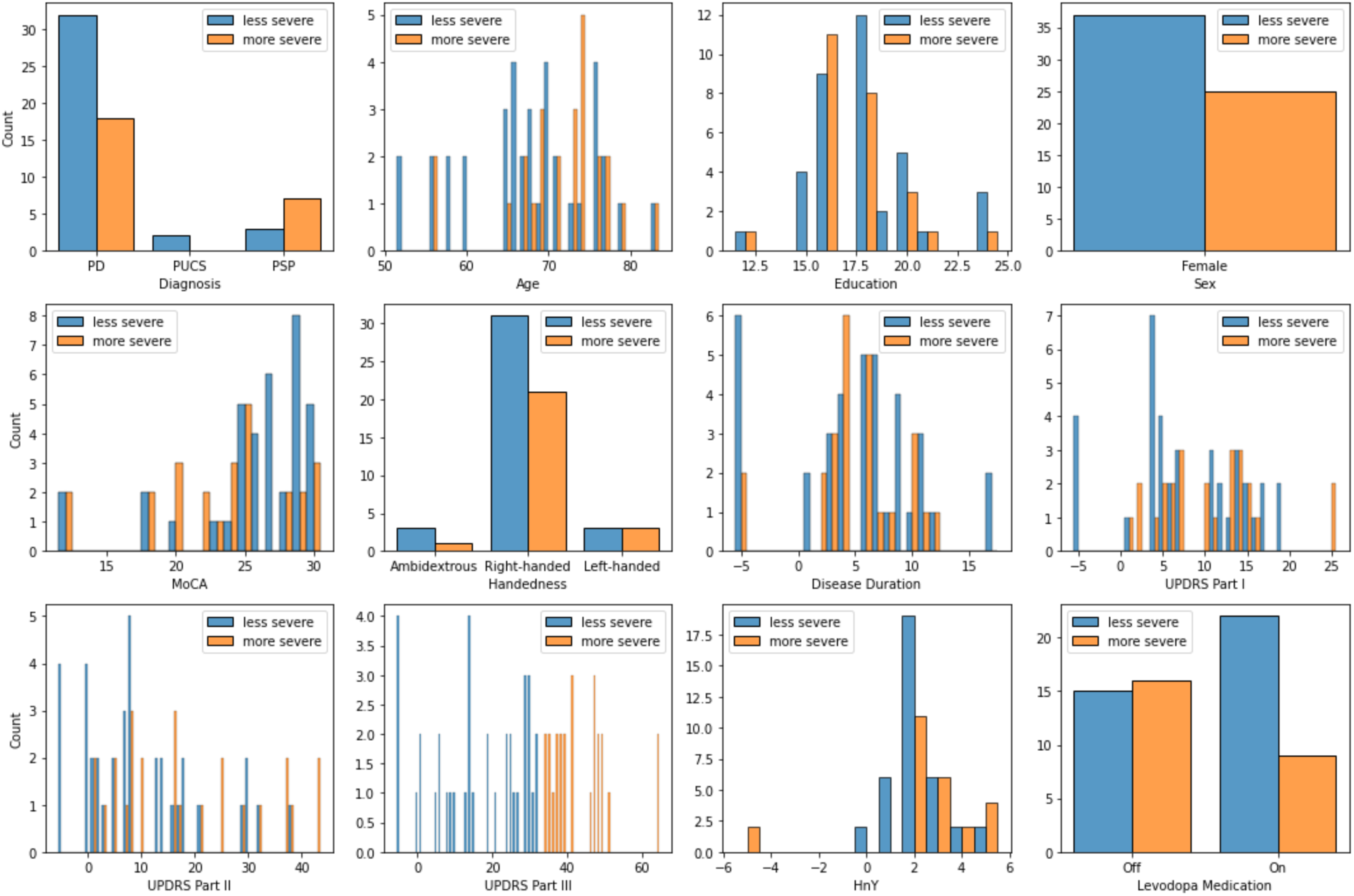
Histograms showing distributions of clinical and demographic characteristics between low and high motor symptom severity groups.

**Supplemental Figure 2:**
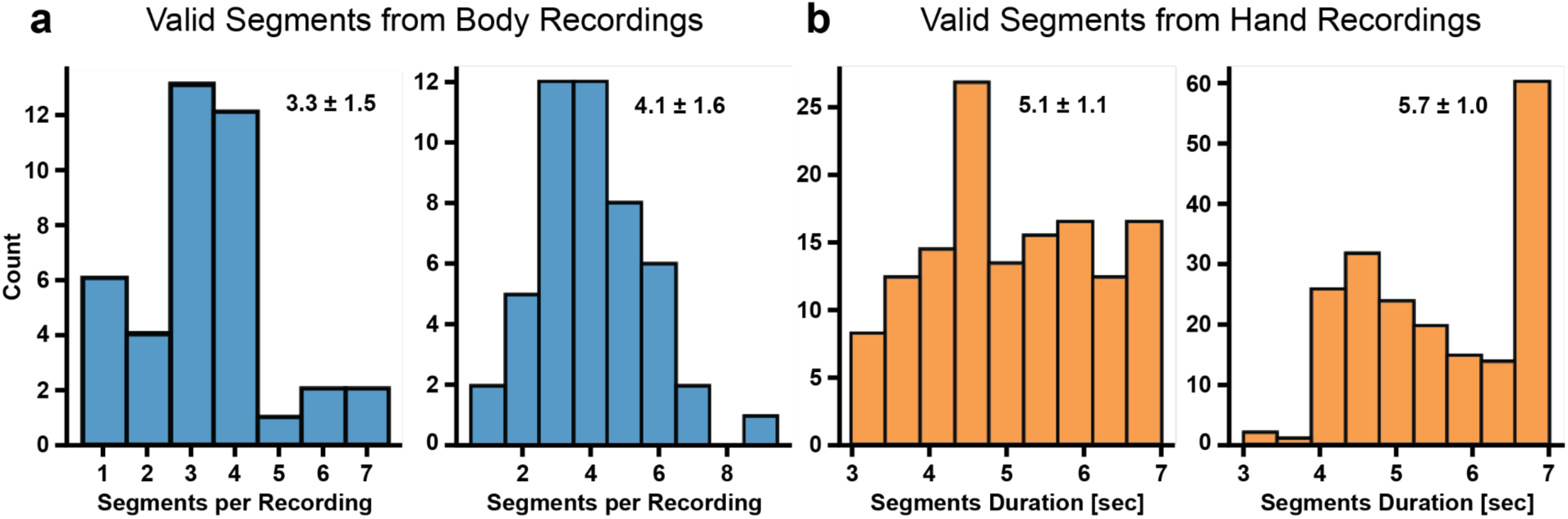
Histograms showing time segment counts per recording and their duration.

**Supplemental Figure 3:**
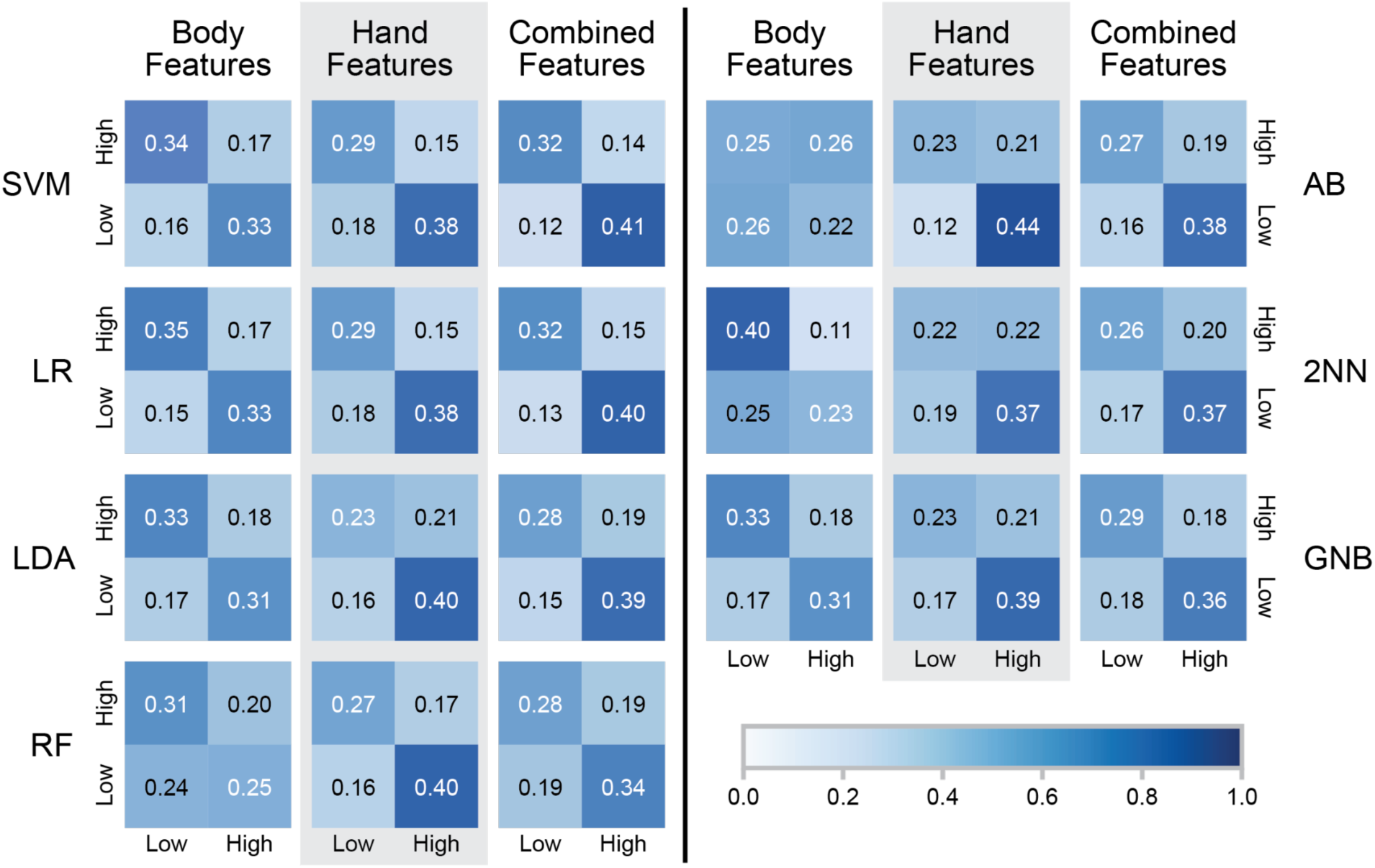
Average confusion matrices of selected classifiers. LDA = linear discriminant analysis; LR = logistic regression; SVM = support vector machine; RF = random forest; AB = adaptive-boosted trees. Compared to classifiers trained with gait features, classifiers trained with finger-tapping features misclassified the low motor impairment class more. Classifiers trained with both gait and finger-tapping features had more confidence and accuracy when predicting the motor impairment labels.

**Supplemental Figure 4:**
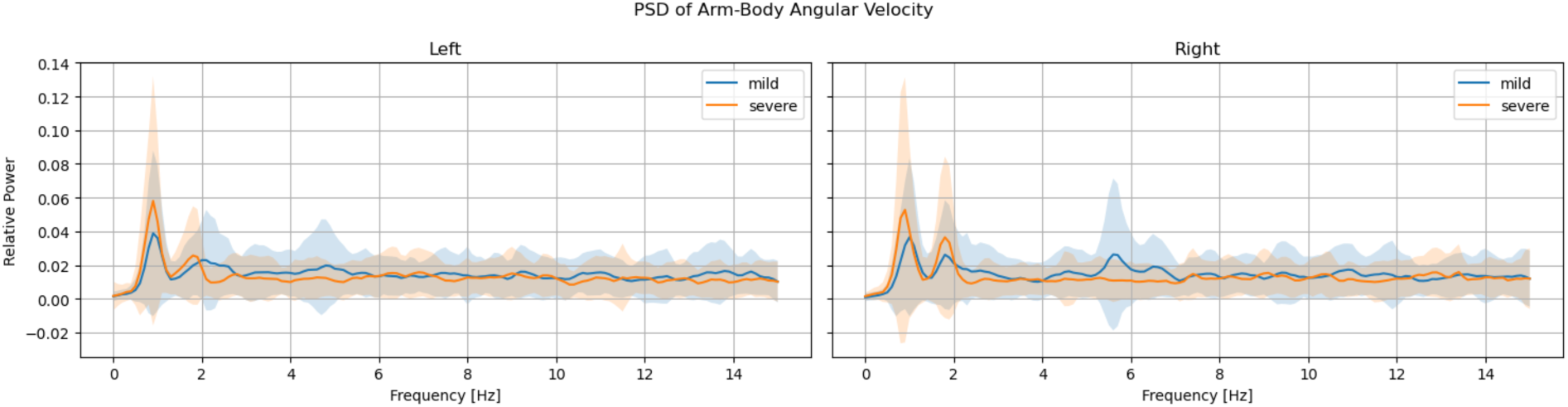
Average relative PSDs of arm-body lateral angular velocity of all participants. The shaded areas correspond to ± 1 standard deviation.

https://drive.google.com/file/d/1yZmQd2WJKd255fFlKta8yG0YzLzgkF-R/view?usp=share_link

https://drive.google.com/file/d/1_rsVCPP1qmm3Bei47KWrx0mR1pPP9ge7/view?usp=share_link

**Supplemental Video 1: Visualizations of extracted landmark kinematics of example pose and hand recordings.** At any point in the videos, a green background indicates that the given frame is error-free and usable, whereas a red background indicates that the frame is to be discarded.

## Notes

### Competing Interest Statement

The authors have declared no competing interest.

### Author Declarations

Privacy and confidentiality protection for this study have been explicitly addressed with the University of California, San Francisco IRB approval. The raw videos were not authorized for release.

